# Deriving Explainable Metrics of Left Ventricular Flow by Reduced-Order Modeling and Classification

**DOI:** 10.1101/2023.10.03.23296524

**Authors:** María Guadalupe Borja, Pablo Martinez-Legazpi, Cathleen Nguyen, Oscar Flores, Andrew M. Kahn, Javier Bermejo, Juan C. del Álamo

## Abstract

**Background:** Extracting explainable flow metrics is a bottleneck to the clinical translation of advanced cardiac flow imaging modalities. We hypothesized that reduced-order models (ROMs) of intraventricular flow are a suitable strategy for deriving simple and interpretable clinical metrics suitable for further assessments. Combined with machine learning (ML) flow-based ROMs could provide new insight to help diagnose and risk-stratify patients.

**Methods:** We analyzed 2D color-Doppler echocardiograms of 81 non-ischemic dilated cardiomyopathy (DCM) patients, 51 hypertrophic cardiomyopathy (HCM) patients, and 77 normal volunteers (Control). We applied proper orthogonal decomposition (POD) to build patient-specific and cohort-specific ROMs of LV flow. Each ROM aggregates a low number of components representing a spatially dependent velocity map modulated along the cardiac cycle by a time-dependent coefficient. We tested three classifiers using deliberately simple ML analyses of these ROMs with varying supervision levels. In supervised models, hyperparameter gridsearch was used to derive the ROMs that maximize classification power. The classifiers were blinded to LV chamber geometry and function. We ran vector flow mapping on the color-Doppler sequences to help visualize flow patterns and interpret the ML results.

**Results:** POD-based ROMs stably represented each cohort through 10-fold cross-validation. The principal POD mode captured >80% of the flow kinetic energy (KE) in all cohorts and represented the LV filling/emptying jets. Mode 2 represented the diastolic vortex and its KE contribution ranged from <1% (HCM) to 13% (DCM). Semi-unsupervised classification using patient-specific ROMs revealed that the KE ratio of these two principal modes, the vortex-to-jet (V2J) energy ratio, is a simple, interpretable metric that discriminates DCM, HCM, and Control patients. Receiver operating characteristic curves using V2J as classifier had areas under the curve of 0.81, 0.91, and 0.95 for distinguishing HCM vs. Control, DCM vs. Control, and DCM vs. HCM, respectively.

**Conclusions:** Modal decomposition of cardiac flow can be used to create ROMs of normal and pathological flow patterns, uncovering simple interpretable flow metrics with power to discriminate disease states, and particularly suitable for further processing using ML.

## 1. INTRODUCTION

Current functional hemodynamic imaging relies on the laws of fluid dynamics, or their simplifications, applied to intracardiac blood flow data (Bermejo et al., 2015; Mele et al., 2019). Studies have shown the potential of flow-derived metrics to improve the characterization of systolic (Yotti et al., 2014) and diastolic function (Little et al., 1995), and to determine the risk of intracardiac thrombosis (Harfi et al., 2017). Left ventricular (LV) flow analyses are also useful to detect subclinical LV dysfunction (Eriksson et al., 2013), predict functional capacity in patients with cardiomyopathy (Stoll et al., 2019), and calculate cardiac hemodynamic forces, which may help to detect subclinical myocardial dysfunction (Vallelonga et al., 2021). However, translation of flow-related metrics based on imaging into clinical practice has been slower than the technological development of imaging technology.

A challenge for establishing flow-derived biomarkers is the choice of a specific metric or a combination of metrics (e.g., vortex circulation, kinetic energy, etc.). This choice can be straightforward for conditions with a well understood pathophysiology clearly linked to hemodynamics. For instance, blood stasis is an essential factor in thrombogenesis (Watson et al., 2009), and quantifying LV stasis offers routes to translation (Martinez-Legazpi et al., 2018; Rodríguez-González et al., 2023). However, when the link between flow and disease is less well understood, as its relationship with systolic function (Watanabe et al., 2008), the low metrics that can diagnose or risk-stratify subjects are more elusive. Of note, there are no systematic procedures for choosing among the many available metrics or deriving new ones.

Modal decomposition, a family of techniques aimed to extract lower-dimensional representations of complex data, has proven effective for drawing out physically important flow features, known as modes, in complex flows (Brunton et al., 2020). The dominant modes can be used to develop reduced-order models (ROMs) that replicate the dynamic behavior of the flow, allowing for simplified simulation and analysis. Among the modal decomposition methods available, proper orthogonal decomposition (POD), also known as principal component analysis, is well established and reproduces most of the flow’s kinetic energy with a reduced number of modes (Taira et al., 2017).

This study is based on two working hypotheses. First, we postulate that POD is an efficient method to reduce the dimensionality of intracardiac flows into simple interpretable metrics. In this context, interpretability is understood as knowledge of the specific vector flow pattern(s) associated to each metric. Second, we submit that these metrics can be readily combined with straightforward ML models for clinical exploitation as in simple binary classification problems (e.g., healthy vs. diseased).

To test these hypotheses, we used POD on clinical 2D color-Doppler echocardiograms to derive ROMs of LV flow that accurately discriminated subjects from three cohorts: healthy individuals, patients with dilated cardiomyopathy, and patients with hypertrophic cardiomyopathy. Balancing ROM simplicity with classification accuracy provides a systematic approach to derive novel LV flow metrics whose interpretation is facilitated by enhancing Doppler data by vector flow mapping. The favorable classification accuracy, obtained with overtly uninvolved machine learning methods and using clinically accessible input data, suggests a novel strategy to realize the untapped potential of non-invasive cardiac flow imaging. Further research is needed to validate these findings and to investigate the utility of flow-based biomarkers in more complex clinical problems.

## 2. METHODS

### 2.1 STUDY COHORTS

We prospectively selected 209 subjects from an existing database of previous studies (Benito et al., 2019; Bermejo et al., 2014; Martinez-Legazpi et al., 2014), including 81 patients with non-ischemic dilated cardiomyopathy (DCM), 51 patients with hypertrophic cardiomyopathy (HCM), and 77 normal volunteers who served as control group (Control). Inclusion criteria for all participants were: 1) presence of sinus rhythm; 2) suitable apical ultrasonic window and 3) absence of significant aortic regurgitation (grade < 2-3). All HCM and DCM patients were included after robust diagnosis (based on clinical, imaging, familial, and genetic data), had angiographically proven absence of significant coronary artery disease, and were clinically stable at the time of imaging. The control group was selected by trying to match the DCM cohort by age and its inclusion criteria were: 1) absence of known or suspected cardiovascular disease, 2) normal electrocardiographic and echocardiographic Doppler examinations, and 3) no history of diabetes or hypertension. The Institutional Ethics Committee at Hospital Gregorio Marañón approved the study, and all participants provided written informed consent.

### 2.2 IMAGE ACQUISITION

Echocardiographic examinations were performed using a Vivid 7 scanner and broadband transducers (GE Healthcare). Conventional echocardiographic data were recorded and measured following current recommendations (Lang et al., 2015). Two-dimensional (2D) sequences were obtained from parasternal and apical views ensuring complete apical visualization without foreshortening and used to measure left-ventricle (LV) volumes and ejection fraction. Pulsed-wave Doppler spectrograms were obtained at the level of the mitral tips and the LV outflow tract. The temporal features of the cardiac cycle were measured from spectral Doppler recordings using EchoPac software (version 110.1.2, GE Healthcare) and stored to serve as temporal landmarks in postprocessing. In addition, color-Doppler images encompassing the whole LV chamber were obtained in the apical three-chamber view followed by 2D cine-loops for flow field quantification (∼10 cycles, frame rate > 60 Hz).

### 2.3 VECTOR FLOW MAPPING

To enhance the interpretability of the reduced-order models and associated classifiers, we derived time-resolved 2D blood velocity fields from color-Doppler data inside the LV using vector flow mapping (VFM), an algorithm thoroughly described in previous studies (Avesani et al., 2021; Garcia et al., 2010; Hvid et al., 2023; Minami et al., 2021). The VFM algorithm is fed by a color-Doppler acquisition and integrates the continuity equation imposing no-penetration boundary conditions at the LV endocardium segmented from the 2D cine-loops. This provides the crossbeam flow velocity under the hypothesis of planar flow.

### 2.4 UNIFIED SPATIO-TEMPORAL COORDINATE SYSTEM

To compare LV flow across patients and ensure that classifiers were blinded to chamber geometry and function (e.g., LV volume and ejection fraction), we adopted a unified LV coordinate system that remapped all patients’ flow data into a fixed rectangular domain, as in our previous works(Bermejo et al., 2014). This coordinate system’s origin is the midpoint of the LV base, its y-axis runs from the origin to the apex, and its x-axis runs in the orthogonal direction (**Figure 1**). The spatial coordinates are normalized so y=1 at the apex and x=-/+ 1 at the lateral and septal walls of the LV, respectively. A constant number of mesh points was used in the x and y directions (Nx=25, Ny=40). We also rescaled each patient’s flow into a dimensionless time scale,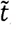, beginning at aortic valve opening,*t*_*AVO*_, to minimize the effect of heart rate variability across patients, i.e.,

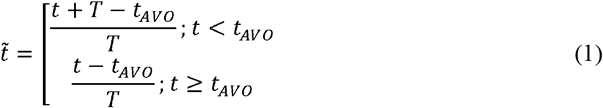

where *T* is the cardiac period of each patient.

**Figure 1:**
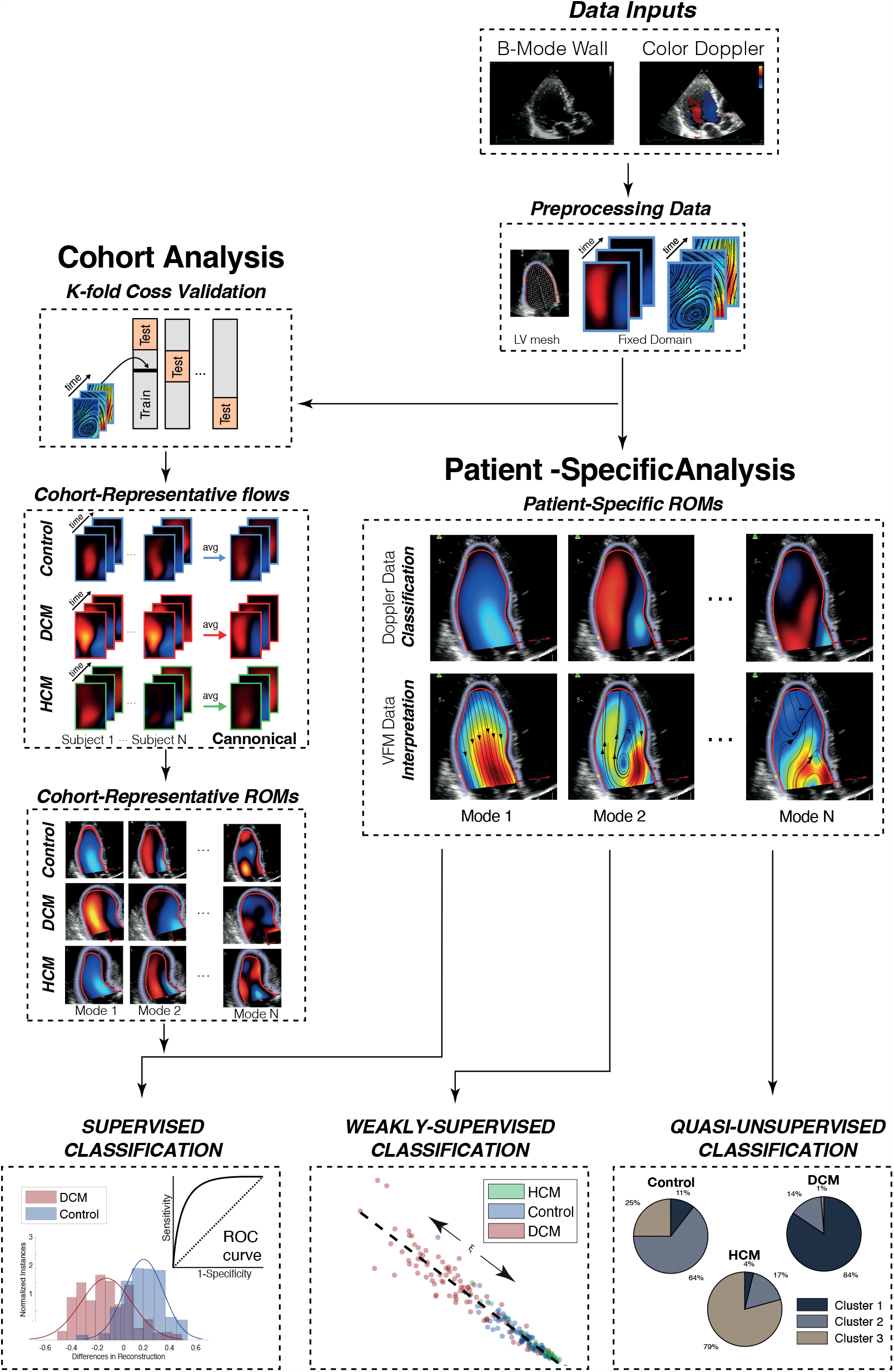
Overview of the methods used for image acquisition and postprocessing.

### 2.5 REDUCED-ORDER MODELING OF LV FLOW

We used the method of snapshots to obtain two common variants of POD: mean-centered (PODmc) and non-mean-centered (PODnmc) (Miranda, 2008). The input velocity data were defined respectively as 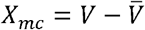 or *X*_nmc_ = *V*, where is a matrix whose columns, *v*_*i*_, represent temporal snapshots of the velocity field (*i* = 1, ., *N*), and 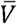is the mean velocity. For color-Doppler fields, *v*_i_ is a vector of length *M* = *N*_*x*_ *N*_*y*_ containing the Doppler velocity in all points of the unified rectangular domain. For 2D velocity fields from VFM, *v*_*i*_ =[*v*_*x*1_, *v*_*x2*_, …, *v*_*xM*_, *v*_*y*1_, *v*_*y2*_, …, *v*_*yM*_] is a vector of length 2M containing the *v*_*x*i_ and *v*_*y*i_ components at the same *M* points. The result of POD is

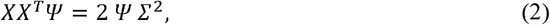

where the columns of Ψ are a set of orthogonal eigenvectors representing flow patterns and Σ^2^ is a diagonal matrix containing the kinetic energy (KE) associated with each eigenvector in PODnmc, or its fluctuations with respect to the mean in PODmc.

#### *Cohort-representative* ROMs were built in two steps

First, we computed cohort-representative flow maps in the unified LV coordinate system by averaging across patients within each group. We then performed POD modes on the resulting flow maps, obtaining a set of eigenmodes ψ ^cohort^, ranked by their KE content. These modes were used to form *cohort-representative* ROMs on which to project a test subject LV flow field, i.e., 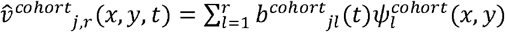 where j denotes the test subject index, *r*< *N* is the number of modes kept in the ROM, and *b*_*jl*_*(t)* is the time-varying contribution of mode *l* to subject’s *j* KE. The accuracy of the ROMs was assessed using the *model residual*, defined as the normalized L2 norm of the difference between the original and the reconstructed flow fields:

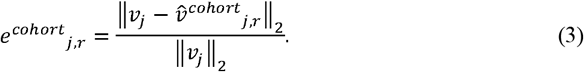

*Patient-specific ROMs* using the POD of each individual patient’s LV flow, *i*.*e*., 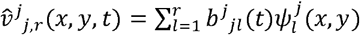. The time-averaged values of the *b*^*j*^_*jl*_*(t)* coefficients are used for classification as described below. **Figure 1** depicts the entire methodology.

### 2.6 SUPERVISED CLASSIFICATION BASED ON COHORT-REPRESENTATIVE REDUCED ORDER MODELS

We built supervised classifiers using the *cohort-representative* ROMs of LV flow, and tested these classifiers on the DCM, HCM, and Control groups. For a given test patient, the classifiers utilized the residuals of alternative cohort-specific ROMs (*i*.*e*.,*e*^*CON*^, *e*^*DCM*^, and *e*^*HCM*^). Equivalently, residual differences with respect to the control case can be used (i.e.,*E*_*DCM -VOL*_ =*e*^*DCM*^*−e*^*VOL*^and *E*_*HCM-VOL*_=*e*^*HCM*^*−e*^*VOL*^). Intuitively, when a subject belongs to a certain cohort, the cohort’s flow ROM reproduces the subject’s flow with lower error than the alternative ROMs (*e*.*g*.,*e*^*DCM*^ <*min(e* ^*VOL*^, *e*^*HCM*^ ) or *E*_*DCM -VOL*_ <*min*(0, *E*_*HCM –VOL*_) for DCM patients). A more accurate, standardized approach was followed, applying linear discriminant analysis (LDA (Abid et al., 2018)) and building receiver operating characteristic (ROC) curves by varying the discrimination cutoff.

#### 2.6.1 K-FOLD CROSS-VALIDATION

To train and validate the classifiers, we split each cohort’s input data into a training and testing dataset using repeated 10-fold cross validation. For each cross-validation run, a set of time-scaled, spatially remapped flow fields was obtained. These flow fields were averaged to obtain a cohort’s canonical flow field, and the canonical field’s POD modes were calculated to build the cohort’s reduced-order model (ROM) for that cross-validation run (**Figure 1-Cohort Analysis**). Classification was performed on the corresponding testing datasets using the trained each cohort-representative ROMs. Statistics gathered from repeating these runs were used to tune model hyperparameters and evaluate the classification performance.

#### 2.6.2 HYPERPARAMETER GRID SEARCH

The classifier based on cohort-specific ROM residuals requires choosing the number of ROM modes, *r*, a methodological hyperparameter. To this end, we conducted hyperparameter grid search (Agrawal, 2020), using two metrics to quantify each ROM’s classification performance. First, we determined the hyperparameter combinations (*r*_*VOL*_, *r*_*DCM*_, *r*_*HCM*_) producing significant residual differences between a test patient’s cohort and the other two cohorts. *P*-values were calculated using a Wilcoxon signed-rank one-tail test. Hyperparameter combinations that did not yield significant residual differences (*p*-value > 0.025) were considered non-discriminant and were therefore discarded. The second metric was the AUC obtained from LDA decision boundaries, which measures the boundary’s ability to distinguish between cohorts. Specifically, we considered the minimum AUC of the binary classification problems DCM vs. Control, HCM vs. Control, and DCM vs. HCM across all 10-fold CV repeats for each hyperparameter combination. As a bias mitigation strategy, nested cross validation was used when computing the AUC values. To prevent excessive computational cost of unconstrained grid search, we constrained the number of hyperparameter combinations by ignoring ROMs with *r*> 30 and bounding *r*_*VOL*_, *r*_*DCM*_, and *r*_*HCM*_, to be in a ± 5 range of each other. To prioritize simpler models and prevent overfitting, we minimized a metric inspired in Akaike’s information criterion, *AIC* =−*AUC +* 0.01 x (*r*_*VOL*_ + *r*_*DCM*_ + *r*_*HCM*_), which balances increasing AUC by 1% vs. increasing model order by one. **Supplemental Figure 1** summarizes the results of grid search for PODnmc and PODmc ROMs of color-Doppler and vector flow maps, including each case’s three top-ranked hyperparameter combinations.

### 2.7 WEAKLY-SUPERVISED AND QUASI-UNSUPERVISED CLASIFICATION BASED ON PATIENT-SPECIFIC REDUCED ORDER MODELS

In addition to the supervised method described above, we explored whether *patient-specific* ROMs of LV flow contain intrinsic properties (*e*.*g*., their level of sparsity), common across patients of a same cohort, that allowed for classification. A notable advantage of such classifiers is their potential for significantly reducing user supervision. Thus, we built weakly-supervised and quasi-unsupervised classifiers following standard analyses of the point clouds generated by the coefficients,

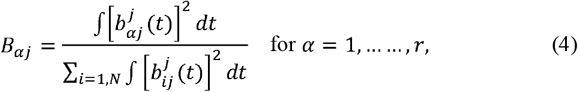

which represent the fraction of the flow KE accounted by the POD mode *α* for each patient *j*.

First, we built *weakly-supervised* classifiers using LDA to calculate decision boundaries between the point clouds created by *B*_*αj*_(*α* = 1, …, *r*) for each cohort. Since the KE content of POD eigenmodes decayed very steeply with mode number (see **Supplemental Figure 2)**, we restricted our analysis to *r*= 2 for the sake of simplicity. Generalizing this procedure to *r*> 2 would be straightforward. ROC AUCs were computed for the binary classification problems DCM vs. Control, DCM vs. HCM, and HCM vs. Control.

Finally, we tested a *quasi-unsupervised* classifier based on k-means clustering (Celebi and Aydin, 2016) of the point clouds created by *B*_*αj*_(*α* = 1, …, *r*) for each cohort. The only supervision provided to this method was the number of clusters, *k = 3*. The results were quite insensitive to the distance metric utilized by k-means, and k-medoids and mixed Gaussians models produced similar results.

### 2.8 STATISTICAL ANALYSES

Descriptive variables are shown as mean and standard deviation, unless otherwise specified. Kruskal-Wallis rank sum test and Fisher’s exact test were used to compare these variables among the HCM, DCM and Control groups. Also, one-way analyses of variance, followed by Tukey contrasts, were used to compare quantitative variables among the different cohorts. ROC curves were calculated on the aggregated binary problems of comparing HCM and DCM, Control and DCM, and Control and HCM. The area under the ROC curve (AUC) was computed to quantify the accuracy of binary classifiers, whereas the multi-class AUC measure, *M* (Hand and Till, 2001), was computed as a measure of multi-class classification performance. Statistical significance was established at the p< 0.05 level. Statistical analyses were performed using R version 3.6.1 and MATLAB (R2020). t-distributed stochastic neighbor embedding was performed on the multiplets formed by normalized eigenvalues coming from patient-specific POD, *B*_*αj*_ (*α*= 1, …, *r*= 15), using MATLAB built-in functions with Euclidean distance.

## 3. RESULTS

### 3.1 CLINICAL DATA AND CONVENTIONAL ECHOCARDIOGRAPHY

Demographic, clinical, and echocardiographic data are shown in **Tables 1 and 2**. Compared to controls, DCM patients were older (62 ± 14 vs. 46 ± 18 years old, p< 0.001) and showed larger indexed LV chamber volumes (151 ± 71 vs. 91 ± 26 mL and 107 ± 67 vs. 34 ± 11 mL for EDV and ESV respectively, p< 0.001) and lower ejection fractions (32 ± 12 vs. 63 ± 5 %, p< 0.001). Compared to controls, HCM patients had smaller indexed LV chamber end diastolic volumes (42 ± 13 vs. 51 ± 12 mlL/m^2^, p= 0.02) and larger ejection fraction (67 ± 7 vs. 63 ± 5 %, p= 0.03). Both cardiomyopathy groups had high E/e’ ratios compared with controls, with values of 11.3 ± 6.1, 10.5 ± 6.51, and 5.7 ± 2.1 for DCM, HCM and Control, respectively (p< 0.001 for both DCM vs. Control and HCM vs. Control).

**TABLE 1:**
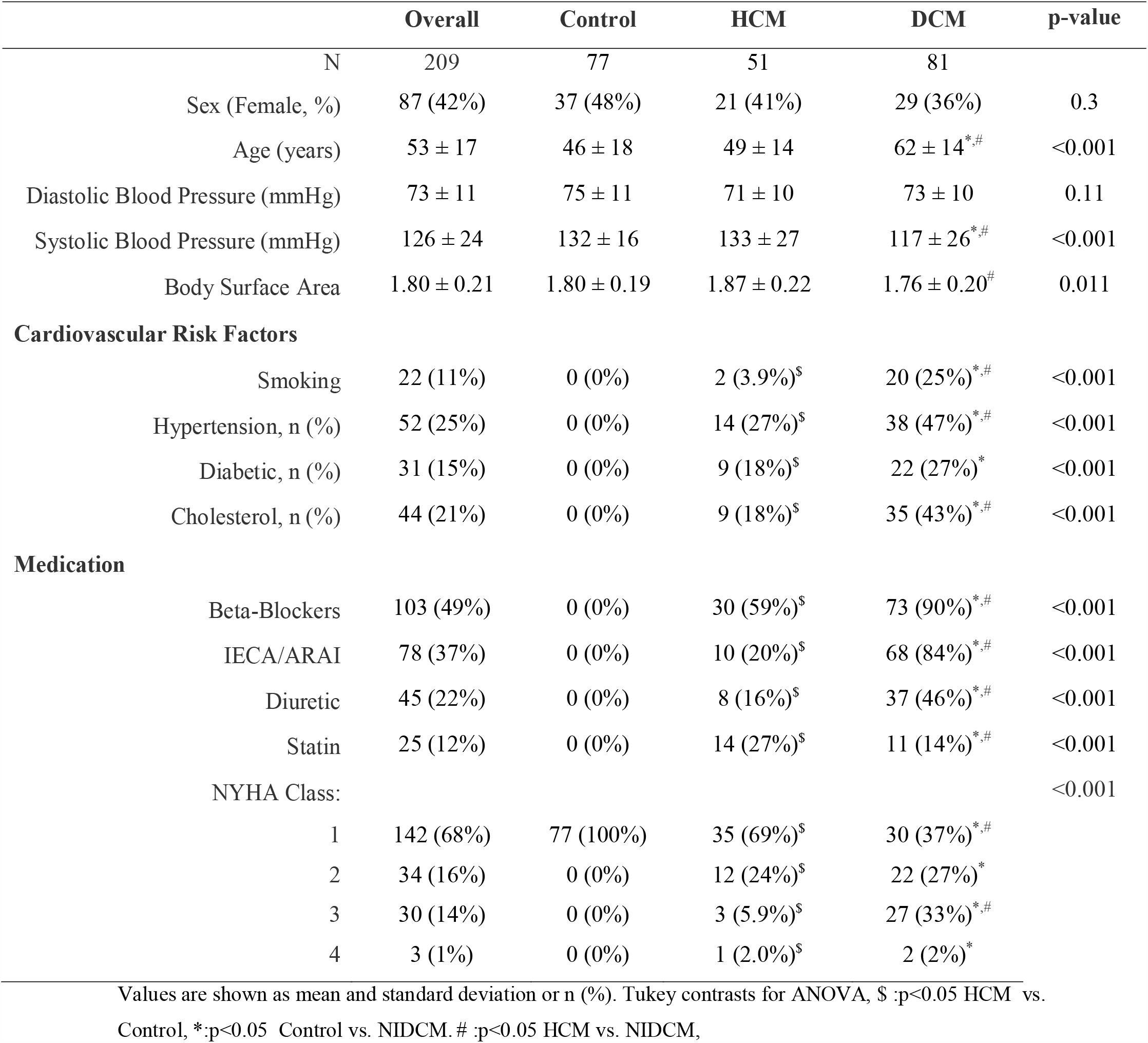
Clinical data.

**TABLE 2:** Echocardiographic Data.

|  | Overall | Control | HCM | DCM | p-value |
| --- | --- | --- | --- | --- | --- |
| LV End Diastolic Volume (mL) | 112 ± 58 | 91 ± 26 | 79 ± 29 | 151 ± 71<sup>*,#</sup> | <0.001 |
| LV End Systolic Volume (mL) | 61 ± 56 | 34 ± 11 | 27 ± 12 | 107 ± 67<sup>*,#</sup> | <0.001 |
| LV Ejection Fraction (%) | 52 ± 18 | 63 ± 5 | 67 ± 7<sup>\$</sup> | 32 ± 12<sup>*,#</sup> | <0.001 |
| LV End Diastolic Volume Index (mL/m<sup>2</sup>) | 52 ± 23 | 51 ± 12 | 42 ± 14<sup>\$</sup> | 90 ± 35<sup>*,#</sup> | <0.001 |
| LV End Systolic Volume Index (mL/m<sup>2</sup>) | 23 ± 20 | 19 ± 5 | 14 ± 6 | 64 ± 33<sup>*,#</sup> | <0.001 |
| Cardiac Output (L/min) | 5.29 ± 3.59 | 4.70 ± 2.22 | 5.15 ± 2.54 | 6.05 ± 5.04 | 0.5 |
| E-wave peak velocity (cm/s) | 0.70 ± 0.20 | 0.72 ± 0.16 | 0.73 ± 0.23 | 0.65 ± 0.21 | 0.02 |
| A-wave peak velocity (cm/s) | 0.63 ± 0.23 | 0.58 ± 0.18 | 0.64 ± 0.29 | 0.68 ± 0.21<sup>*</sup> | 0.004 |
| E-wave Deceleration Time (ms) | 188 ± 61 | 186 ± 45 | 206 ± 69 | 178 ± 68<sup>#</sup> | 0.026 |
| e' velocity (cm/s) | 0.07 ± 0.04 | 0.11 ± 0.03 | 0.05 ± 0.02<sup>\$</sup> | 0.05 ± 0.02<sup>*</sup> | <0.001 |

Values are shown as mean and standard deviation. Tukey contrasts for ANOVA, \$:p< 0.05 HCM vs. Control, \*: p< 0.05 Control vs. NIDCM. #: p< 0.05 HCM vs. NIDCM.

### 3.2 COHORT-REPRESENTATIVE LEFT VENTRICULAR FLOW MAPS

After remeshing all patients flow data, cohort-representative flow maps were obtained by averaging across patients for each 10-fold CV training set. To facilitate interpretation, examples of these from 2D vector flow maps were meshed back into patient-specific anatomies and represented in **Figure 2**. Because of the averaging, some fine-scale features of the patient-specific flow dynamics were smoothed out. For instance, the small early-diastolic inferolateral counterclockwise section of the diastolic vortex ring was absent in the cohort-averaged flow maps of controls. Despite this averaging effect, the cohort-averaged flow maps captured the distinct flow features reported in single normal, DCM, and HCM subjects by different modalities (Bermejo et al., 2014; Elbaz et al., 2014; Pedrizzetti et al., 2014). For example, the cohort-averaged flow displayed a stronger anteroseptal vortex core in DCM compared to control (Bermejo et al., 2014), and this pattern was mostly unobservable in the HCM cohort due to confinement by the abnormally narrow LV morphology (Martinez-Legazpi et al., 2014). These results were robust across all realizations generated via 10-fold cross-validation.

**Figure 2:**
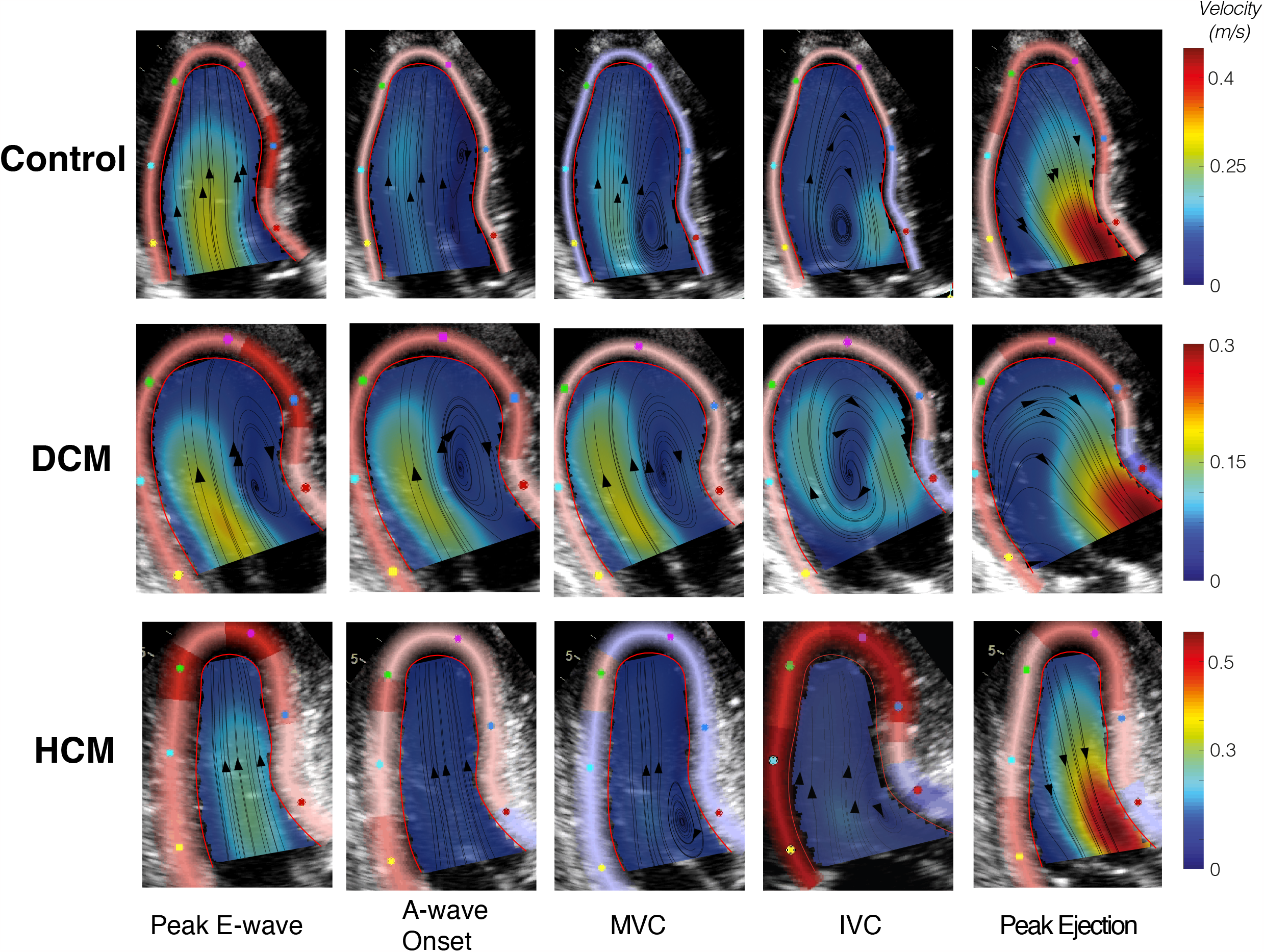
Cohort-representative intraventricular velocity fields from vector flow mapping (VFM). Black lines are instantaneous streamlines and arrows indicate flow direction. Flow speed is indicated by colormaps according to the colorbar on the right-hand side of each row. Patient-specific VFM velocity fields were temporally aligned, mapped onto the rectangular unified spatio-temporal reference system, averaged across the cohort, and then remapped into a patient-specific anatomy randomly chosen from the corresponding cohort. The top, middle, and bottom rows represent the Control, DCM, and HCM cohorts, respectively. Columns represent key events along the cardiac cycle: Peak E-wave, A-wave Onset, Peak A-wave, Mitral valve closure (MVC) and Aortic Valve Opening (AVO).

### 3.3 COHORT-REPRESENTATIVE REDUCED-ORDER MODELS OF LEFT VENTRICULAR FLOW

We applied PODmc to the cohort-averaged color-Doppler maps. **Figure 3** shows results of this analysis for a 10-fold CV example from each cohort, meshed back into patient-specific anatomies. To further aid interpretation, **Figure 4** shows similar results for 2D vector flow maps. In both figures, the four highest ranked modes, ψ _i_ (*x, y), i*, = 1, . .4, are represented, together with the temporal variation of their amplitude across the cardiac cycle, *b*_*i*_ *(t)*. Altogether, these modes accounted for more than 99.5% of the flow’s fluctuating KE.

**Figure 3:**
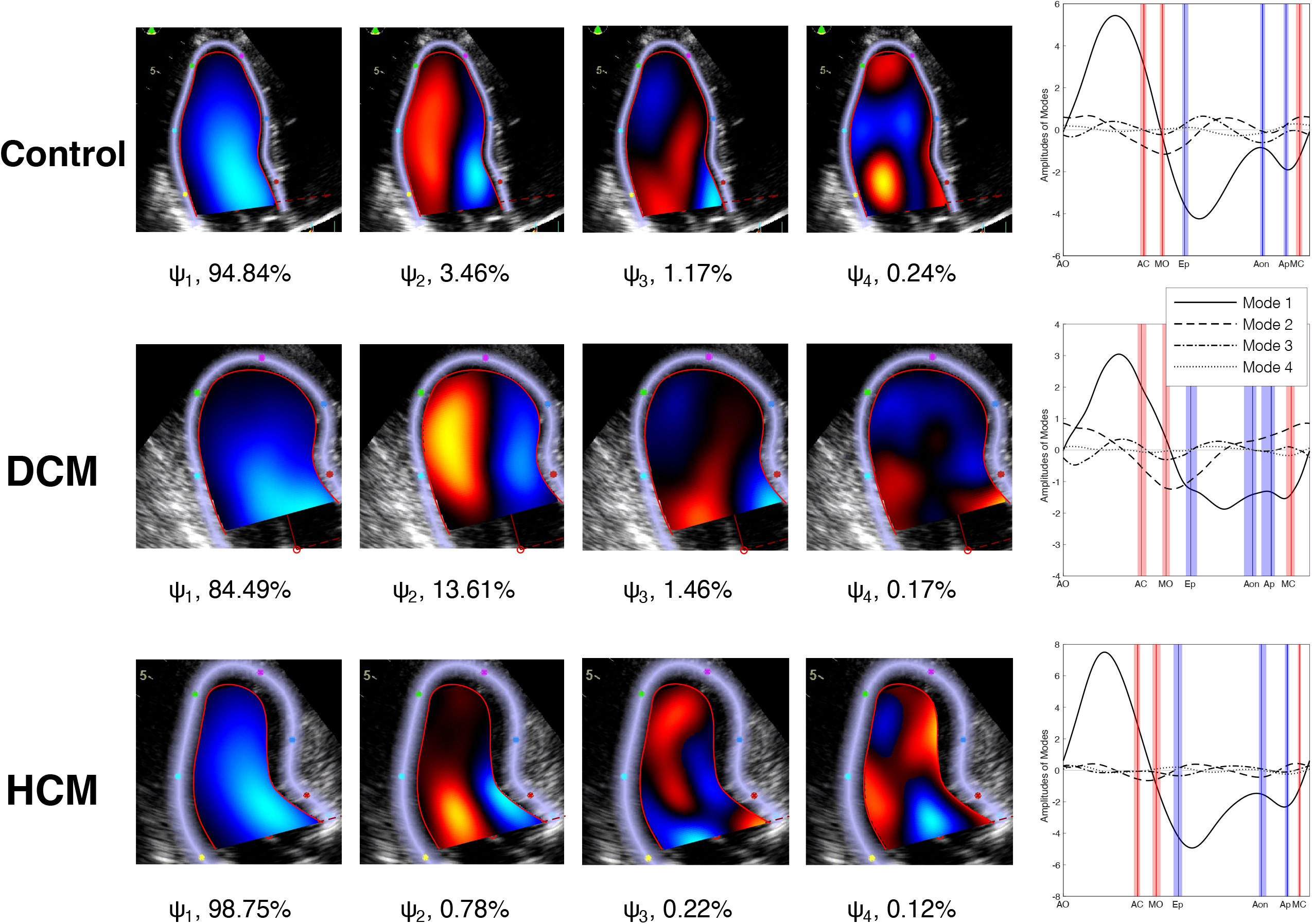
Mean-centered POD of cohort-representative intraventricular color-Doppler flow maps. Color-Doppler fields were mapped onto the rectangular unified spatio-temporal reference system to calculate the POD. Then, the POD results were remapped onto a patient-specific anatomy randomly chosen from the corresponding cohort to facilitate visualization. The top, middle, and bottom rows indicate the Control, DCM and HCM cohorts, respectively. Columns 1^st^ to 4^th^ represent the four highest-ranked POD eigenmode spatial maps 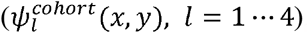, whereas the 5^th^ column depicts these modes’ time-dependent KEs 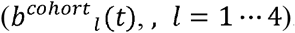. Red/blue lines represent each cohort’s median event times: Aortic Valve Open (AVO), Aortic valve closure (AVC), E-wave onset (MVO), peak E-wave (Ep), A-wave onset (Aon) peak A-wave (Ap) and mitral valve closing (MVC). Shaded red and blue areas are the bootstrap median 95% confidence interval for each cohort.

**Figure 4:**
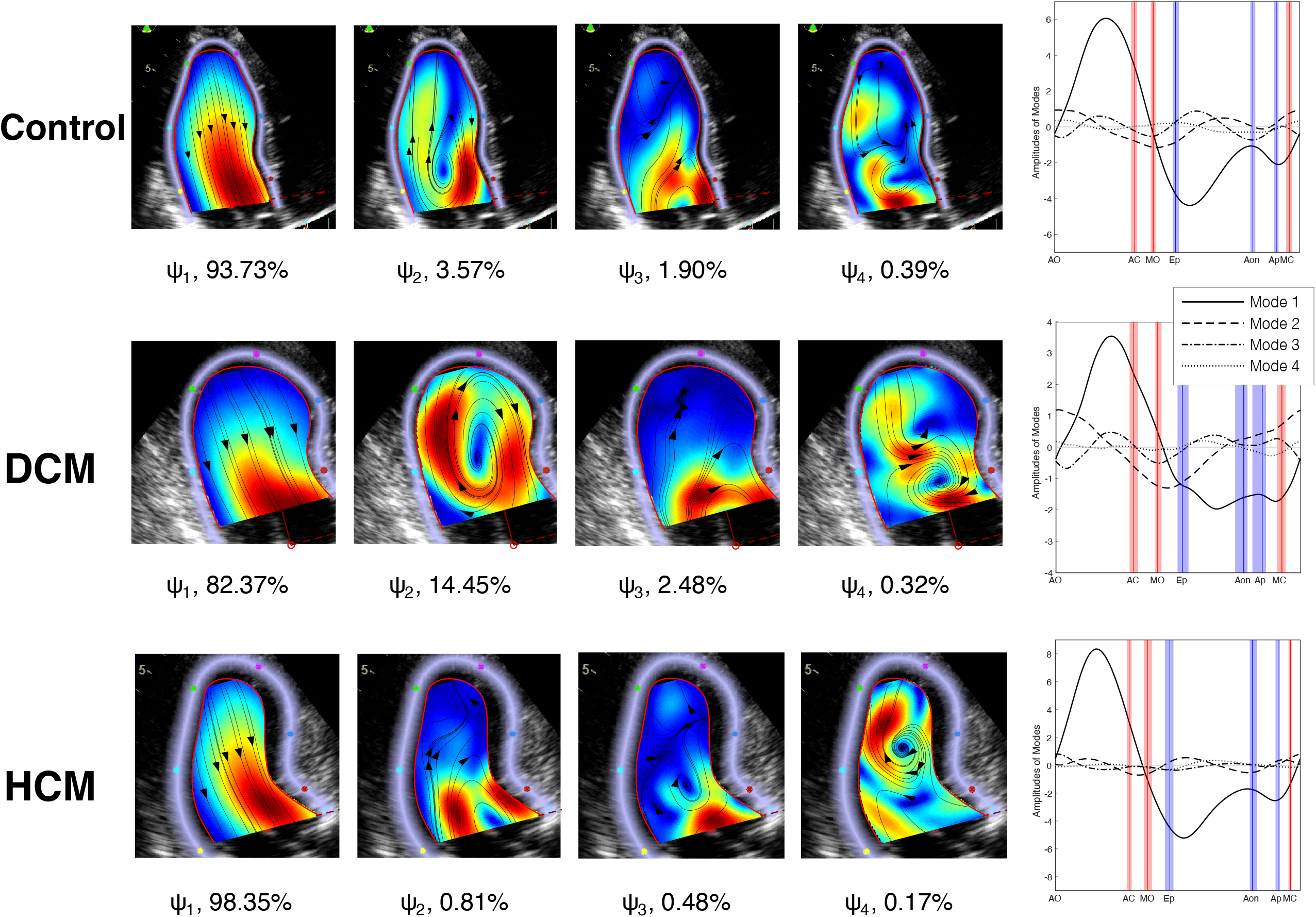
Mean-centered POD of cohort-representative intraventricular vector flow mapping (VFM) velocity fields. VFM fields were mapped onto the rectangular unified spatio-temporal reference system to calculate the POD. Then, the POD results were remapped onto a patient-specific anatomy randomly chosen from the corresponding cohort to facilitate visualization. The top, middle, and bottom rows indicate the Control, DCM and HCM cohorts, respectively. Columns 1^st^ to 4^th^ represent the four highest-ranked POD eigenmode spatial maps 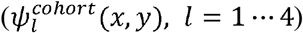, whereas the 5^th^ column depicts these modes’ time-dependent KEs 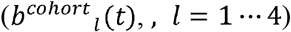. Red/blue lines represent each cohort’s median event times: Aortic Valve Open (AVO), Aortic valve closure (AVC), E-wave onset (MVO), peak E-wave (Ep), A-wave onset (Aon) peak A-wave (Ap) and mitral valve closing (MVC). Shaded red and blue areas are the bootstrap median 95% confidence interval for each cohort.

The highest ranked mode in the three groups was a straight jet, the dominant flow pattern of the filling and ejection phases. Its amplitude reached its maximum positive (base-to-apex flow) value at peak systole, while two negative (apex-to-base flow) amplitude peaks were found at the E and A filling waves. The fraction of fluctuating KE contained in this mode was highly significant although it varied among groups (93-94% in the Control group, 83-85% in DCM, and 98-99% in HCM). The jet pattern formed by mode 1 reached deep into the LV in the Control and HCM groups. However, this pattern was shallower in DCM patients, leaving the apical region disconnected from inflow and outflow jets.

The second mode consisted of a prograde swirling structure near the LV base that linked the inflow and outflow jets. In the healthy cohort, this mode had positive amplitude during E-wave and A-wave deceleration, and early systole. However, its effect was more prominent during early systole than during late filling in DCM patients, whereas it remained weak throughout the cardiac cycle in HCM patients. The contribution of mode 2 to KE fluctuation was largest in the DCM group (13%), followed by the Control group (4%), while it was insignificant in the HCM group (< 1%). Compared to the healthy group, the vortex pattern created by mode 2 was larger and more apically located in DCM patients, whereas it was smaller and more basally located in HCM patients. Modes 3 and 4 represented smaller swirling structures near the LV base and apex, respectively, and accounted for insignificant fractions of the fluctuating KE in all the groups.

Each cohort’s flow medoid, i.e., the patient whose ROM yields the minimal aggregate residual for all other patients in the same cohort, had nearly identical patient-specific PODmc modes as the cohort-representative flow. Furthermore, the non-mean-centered POD analysis produced similar results (**Supplemental Figures 3 & 4**), with the notable exception that the highest-ranked mode in DCM contained a swirling pattern instead of being a straight jet.

Overall, these data are robust and consistent with diastolic vortices being stronger than normal in DCM and weaker than normal in HCM.

### 3.4 SUPERVISED LEFT VENTRICULAR FLOW CLASSIFICATION BASED ON COHORT-REPRESENTATIVE REDUCED-ORDER MODELS

Grid search analysis consistently ranked (*r*_*VOL*_, *r*_*DCM*_, *r*_*HCM*_)= (1,1,1) as the optimal number of modes for PODmc and PODnmc using either color-Doppler or vector flow mapping data as inputs for classification. Even with only one mode, the PODmc ROMs provided skewed histograms of the residual differences, *E*_*ij*_, leading to favorable AUCs for binary classification between cohorts (**Figure 5**). The DCM vs. HCM classifier performed best, with 0.88 < AUC < 0.93 (bounds of 10-repeated 10-fold CV), followed by the Control vs. DCM classifier, which achieved 0.81 < AUC < 0.86. Lastly, the HCM vs. Control classifier had AUCs between 0.75 and 0.78. The PODnmc ROMs performed slightly better, especially for the HCM vs. Control classification problem (see **Supplementary Figure 5**). Overall, these data indicate that remarkably simple ROMs trained on a given cohort reconstructed patient-specific LV flow maps from that cohort significantly better than alternative cohort ROMs, allowing for deriving simple LV flow metrics that identify disease conditions. The results were robust with respect to the type of modal decomposition (i.e., PODmc vs. PODnmc) and post-processing of color-Doppler images into vector flow maps was not required to achieve accurate classification.

**Figure 5:**
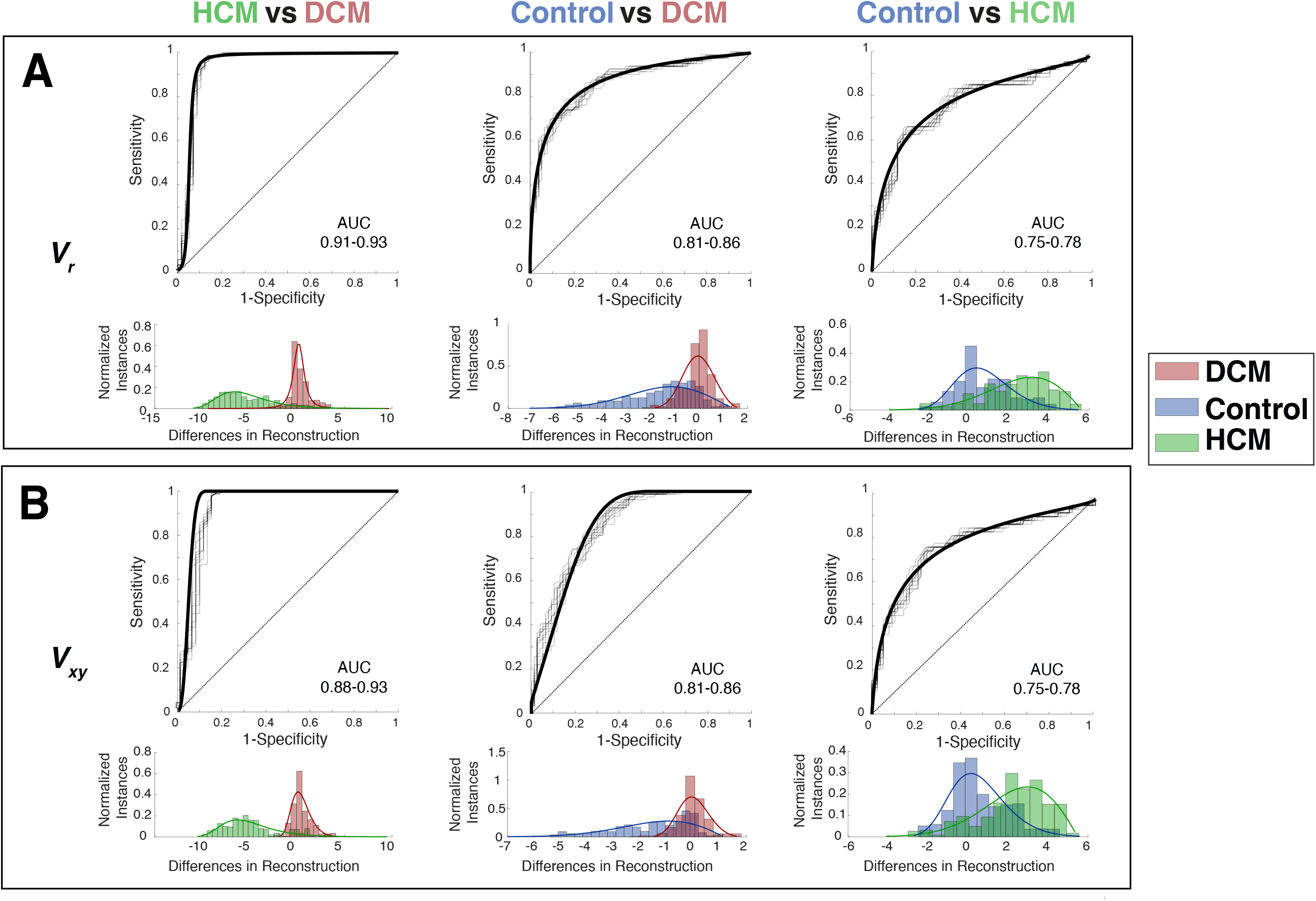
Performance supervised classification based on the residuals of cohort-representative reduced-order models of LV using mean-centered POD. **Panel A**: Model performance using color-Doppler maps as input data. **Panel B**: Model performance using VFM velocity maps as input data. From left to right, each column shows ROC curves (based on approximated distributions and repeated k-fold CV calculations) and residual histograms for the binary comparisons between HCM (green) & DCM (red), Control (blue) & DCM, and Control & DCM.

### 3.5 WEAKLY SUPERVISED LEFT VENTRICULAR FLOW CLASSIFICATION BASED ON PATIENT-SPECIFIC REDUCED-ORDER MODELS

We calculated *patient-specific* PODmc ROMs and the coefficients *B*_*αj*_ representing the fraction of the flow KE fluctuations accounted by mode a for each patient. For simplicity, we restricted our analysis to the two highest-ranked modes, which was justified since these modes contained >97% of fluctuating KE in the *cohort-representative* ROMs, for all three cohorts and for both the color-Doppler and vector flow maps (**Figures 3-4, Supplemental Figure 2**). **Figure 6** shows a scatter plot of *B*_1*j*_and *B*_2*j*_ for all the patients in the three cohorts calculated from Doppler velocity. Similar results were achieved when using VFM velocity fields (**Supplemental Figure 6**). These data were distributed narrowly around a line of negative slope (*B*_2*j*_= -0.63 *B*_1*j*_+ 61, revealing a strong negative correlation (R^2^= 0.85) that allowed for further reducing the model’s dimensionality to one. Moreover, the data clouds corresponding to each cohort exhibited appreciable separation, offering potential for accurate classification. Thus, we used the linear fit to the data points as the discriminant axis, and the distance along this axis from its *B*_*2j*_ *=0* intercept, ξ, as discrimination variable. To confirm this dimensionality reduction approach, we plotted t-distributed stochastic neighbor embedding (t-SNE) maps considering 15 dimensions, i.e., *B* _*αj*_ (*α*= 1, …, *r*= 15), obtaining representations that were similar to the *B*_2*j*_ vs. *B*_1*j*_ scatter plots of **Figure 6** (**see Supplemental Figure 7**).

**Figure 6:**
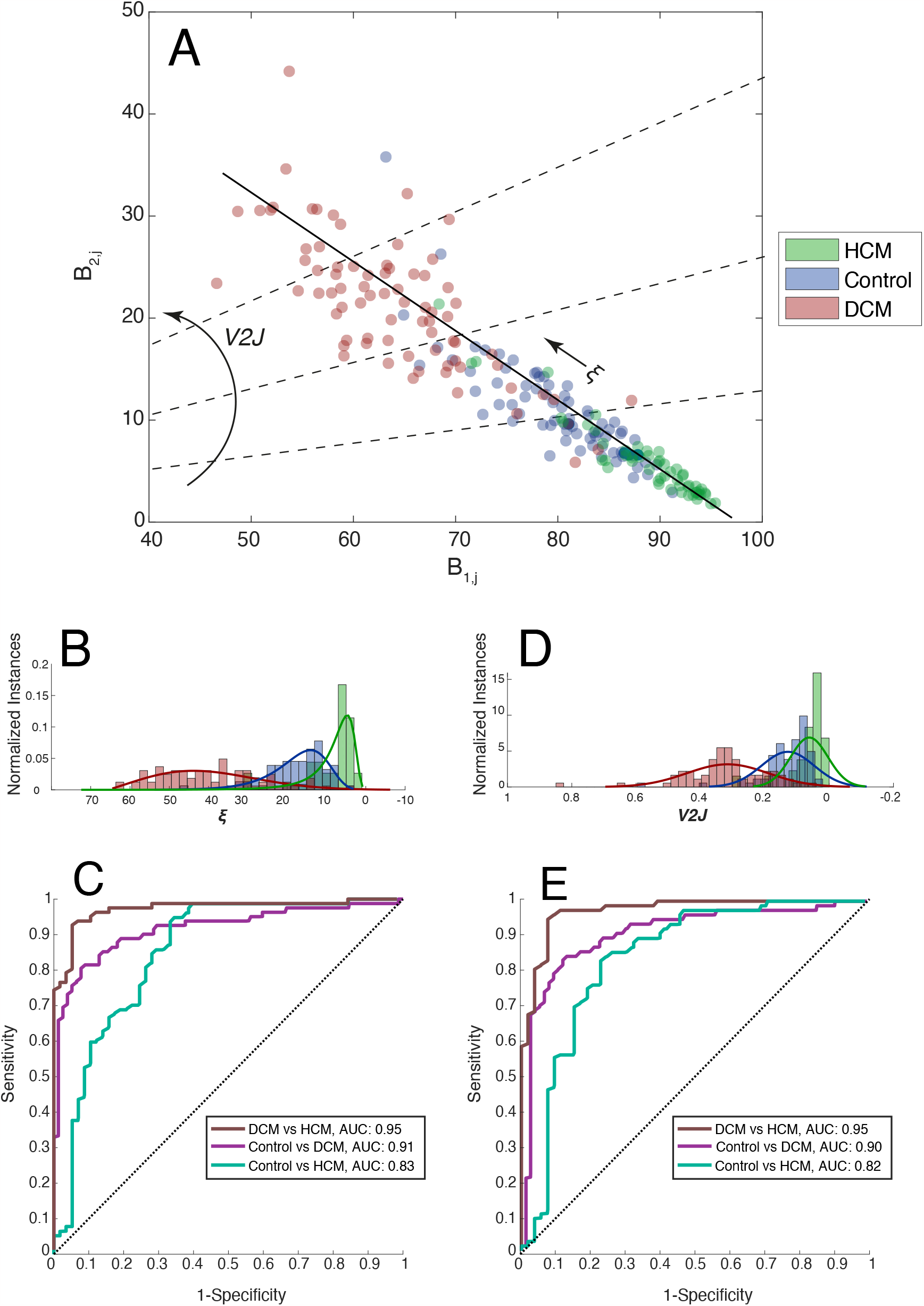
Performance of weakly-supervised classifier based on patient-specific mean-centered POD using color-Doppler velocity fields as input data. **Panel A**: Scatter plot and linear regression of *B*_lj_ and *B*_2j_; each study subject is a data point colored according to their cohort. The dashed lines represent the direction of variation of *V2J* and ξ. **Panels B & D**: Histograms of distance along the discriminant axis ξ and *V2J* respectively for each cohort. **Panels C & E:** Model performance using ξ and *V2J* respectively. ROC curves are shown for the binary classifications: 1) Brown line: HCM (green) vs. DCM (red), 2) Purple line: Control (blue) vs. DCM and 3) Dark green: Control vs. DMC.

To interpret the meaning of the metric ξ, we note that *B*_2_ =1 − Δ*B*−*B*_1_by means of eq. 4, where Δ *B* = Σ_*α*=3,*N*_ *B*_*α*_. This makes ξ interchangeable with the ratio *V*2*J* = *B*_*2*_ / *B*_1_ representing the flow KE contained in the diastolic vortex normalized with the flow KE in the inflow/outflow jets. We named this parameter the *vortex-to-jet ratio* and included it in **Figure 6**. The equivalence between *V2J* and ξ is established by finding the intersection between the empirical fit to *B*_2_ v. *B*_1_ and a series of rays of slope *V2J* emanating from the origin. Assuming Δ *B* << 1, which is reasonable given the steep decay of mode energy with mode rank in our POD models (**Supplemental Figure 2**), this operation results in the equivalence 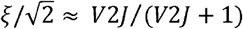

The three patient groups had distinct ξ and *V2J*-histograms (**Figure 6-Panels B and D)**. In the HCM group, both metrics were narrowly distributed near zero, whereas they adopted intermediate values for the Control group. Finally, they reached significantly higher values for the DCM group. We built ROCs for the binary classification problems using ξ and *V2J* as cutoff variables (**Figure 6 – Panels C and E)** and computed the corresponding AUCs for binary classifications and the M metric for three-way classification. The results were highly favorable and rather insensitive to using Doppler or VFM data as inputs (**Table 3**). Of note, the V2J metric computed from color-Doppler maps yielded AUCs of 0.95, 0.90, and 0.82 with 95% confidence intervals (CI) of 0.92-0.97, 0.82-0.94, 0.73-0.88 for discriminating DCM vs. HCM, DCM vs. Control, and HCM vs. Control, respectively. Similar classifiers were tested in patient-specific PODnmc ROMs, obtaining less favorable performance [e.g., V2J -based AUCs from color-Doppler of 0.92 (95% CI: 0.85-0.95), 0.63 (0.53-0.71), and 0.89 (0.83-0.93) for DCM vs. HCM, DCM vs. Control, and HCM vs. Control; see **Table 3** for more details].

**TABLE 3.** Accuracy of weakly supervised classifiers based on patient-specific POD of LV flow fields.

| METRIC |  |  | DCM vs HCM<br>AUC (95% CI) | DCM vs Control<br>AUC (95% CI) | HCM vs Control<br>AUC (95% CI) | 3-way<br>M (95% CI) |
| --- | --- | --- | --- | --- | --- | --- |
| PODmc | $\xi$ | Doppler | 0.96 (0.93-0.98) | 0.91 (0.84-0.94) | 0.83 (0.74-0.90) | 0.91 (0.87-0.94) |
|  |  | VFM | 0.96 (0.94-0.98) | 0.91 (0.86-0.95) | 0.84 (0.74-0.89) | 0.91 (0.87-0.94) |
|  | V2J | Doppler | 0.95 (0.92-0.97) | 0.90 (0.82-0.94) | 0.82 (0.72-0.88) | 0.89 (0.86-0.93) |
|  |  | VFM | 0.96 (0.92-0.97) | 0.90 (0.85-0.94) | 0.82 (0.73-0.88) | 0.90 (0.86-0.93) |
| PODnmc | $\xi$ | Doppler | 0.92 (0.85-0.95) | 0.65 (0.55-0.73) | 0.89 (0.81-0.93) | 0.82 (0.77-0.85) |
|  |  | VFM | 0.93 (0.86-0.96) | 0.64 (0.54-0.72) | 0.90 (0.83-0.94) | 0.82 (0.78-0.86) |
|  | V2J | Doppler | 0.92 (0.85-0.95) | 0.63 (0.53-0.71) | 0.89 (0.83-0.93) | 0.81 (0.77-0.85) |
|  |  | VFM | 0.93 (0.86-0.96) | 0.62 (0.52-0.71) | 0.90 (0.84-0.94) | 0.82 (0.77-0.85) |

### 3.6 QUASI UNSUPERVISED LEFT VENTRICULAR FLOW CLASSIFICATION BASED ON PATIENT-SPECIFIC REDUCED-ORDER MODELS

We applied naive clustering algorithms to the point clouds created by the coefficients *B* _*αj*_ of the *patient specific* PODmc ROMs of LV flow, as described in the Materials & Methods Section. Of note, the only supervision provided to these algorithms was to specify the number of clusters, *k=3*, to match the number of study cohorts. Efforts to estimate the optimal number of clusters using various approaches (e.g., the gap statistic, the silhouette method, etc.) provided results between *k=2* and *k=4*, with most methods yielding *k=2*. **Figure 7-A** displays scatter plots *B*_1*j*_ and *B*_2*j*_ labeled according to the clusters obtained by k-means clustering for *r*= 4, showing a fair level of correspondence between cluster labels and cohort labels (1-Control, 2-DCM, 3-HCM). Classification accuracy improved slightly with the number of ROM modes, converging for *r*≥4, which is sound as a four-mode ROM contained >99.5% of the KE in the three cohort-representative flows. **Supplemental figure 8** represents t-SNE maps embedding 15 dimensions (*B* _*αj*_, *α* = 1,…, 15), showing a similar level of label concordance. **Figure 7** shows pie charts of the distributions of cluster labels per cohort (**Panel B)** and cohort labels per cluster (**Panel C**). Consonant with the results from the other two classification methods presented above, discrimination between DCM and HCM patients was highly accurate, with only 2% of cluster 1 subjects having HCM and 3% of DCM patients being classified in cluster 3. Likewise, discrimination between HCM and Control was least accurate, with 11% of cluster 3 subjects being Control and 16 % of Control subjects being labeled as cluster 3.

**Figure 7:**
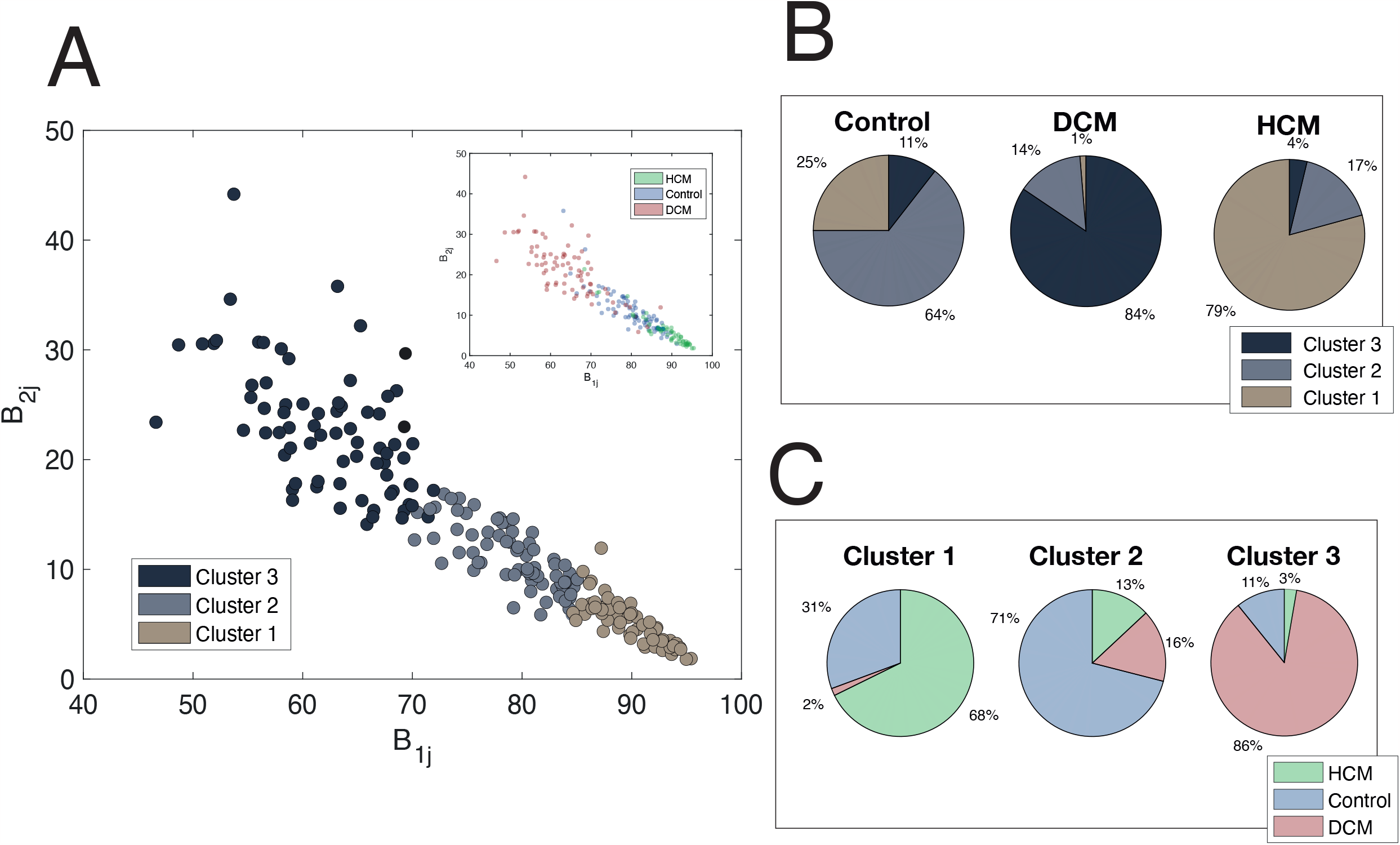
Performance of quasi unsupervised classifier based on patient-specific mean-centered POD using color-Doppler velocity fields as input data. **Panel A**: Scatter plots *B*_lj_ and *B*_2j_ labeled according to the clusters obtained by k-means clustering; each study subject is a data point colored according to their cohort. **Panel B**: Pie charts of the distributions of cluster labels per cohort. **Panel C:** Pie charts of the distribution of cohort labels per cluster.

## 4. DISCUSSION

This study utilized proper orthogonal decomposition (POD) on echocardiographic clinical measurements of left ventricular (LV) flow to create reduced-order models (ROMs) defining the distinctive flow patterns in healthy subjects, patients with dilated cardiomyopathy (DCM), and patients with hypertrophic cardiomyopathy (HCM). We then leveraged these ROMs to derive simple, flow-based metrics that discriminate among study groups using machine learning (ML). To demonstrate the approach and its robustness, we used various well-established ML models to analyze LV flow ROMs in the three groups. We found that a new one-dimensional metric, loosely interpretable as the ratio between the kinetic energies of the transvalvular jets and the diastolic vortex, offers significant discrimination accuracy. The metric is obtained from POD of 2D color-Doppler echocardiograms, a straightforward calculation requiring no assumptions and little expert supervision.

The main LV flow pattern, in addition to the mitral and aortic jets, is a vortex ring formed during diastole (Kilner et al., 2000). This structure has been proposed to facilitate LV filling, conserve inflow kinetic energy (KE), and transfer this energy to the ejection volume (Charonko et al., 2013; Martinez-Legazpi et al., 2014; Pedrizzetti and Domenichini, 2005). Vortex size and KE are altered in cardiomyopathies, augmenting in DCM and almost vanishing in HCM (Bermejo et al., 2014; Elbaz et al., 2014; Martinez-Legazpi et al., 2014). LV flow patterns define the pathlines of blood particles, partitioning the chamber into four regions with distinct transport dynamics (direct flow, retained inflow, delayed ejection, and residual volume) (Eriksson et al., 2013). These regions are altered in cardiomyopathies, most notably the residual volume containing blood with residence time higher than two cycles. While the impacts of LV flow patterns and pathlines in cardiac mechanical energy balance have been a subject of debate (Elbaz et al., 2017; Watanabe et al., 2008), there is consensus of their potential as imaging biomarkers of cardiac health. This potential has stimulated significant developments in flow imaging, such as 4D flow cardiac magnetic resonance (Dyverfeldt et al., 2015), or echocardiographic vector flow mapping (VFM) (Avesani et al., 2021; Garcia et al., 2010) and blood speckle tracking (Daae et al., 2021). However, the significant intra- and inter-patient variability of LV flow dynamics (Bermejo et al., 2014; Sundin et al., 2020) make it difficult to define the normal pattern by narrowly constrained numerical metrics, or quantifying how these metrics are altered in different cardiomyopathies. Therefore, the wealth of information available from the advanced imaging of LV complex flow patterns is currently underutilized in clinical decision support systems.

Modal decomposition techniques have been widely used to study aerospace and industrial flow problems (Taira et al., 2017). Recent studies have applied these techniques to derive ROMs of intraventricular flow in mice (Groun et al., 2022), analyze how dysfunctional heart valves alter hemodynamics in vitro (Darwish et al., 2021; Di Labbio and Kadem), or model the flow in aneurysms (Norouzi et al., 2021; Yu and Durgesh, 2022). Flow classifiers based on ROMs have been proposed to identify distinct regimes in complex fluid problems like stratified turbulence (Ohh and Spedding, 2022). In parallel, the applications of ML to medical imaging analysis and patient phenotyping have grown exponentially in the past decade (Shad et al.; Zhou et al., 2021). The field of echocardiography has not lagged behind. Deep neural networks can be trained to automatically interpret clinical echocardiograms with human-like or super-human accuracy in tasks like heart chamber segmentation, functional assessment, or disease identification (Tromp et al., 2022; Zhang et al., 2018). However, there is a paucity of studies using ML to exploit the multi-dimensional spatio-temporal LV flow datasets currently obtainable from state-of-the-art imaging modalities. This work addresses this paucity by analyzing LV flow color-Doppler maps in an effort to balance data complexity with acquisition burden. Ultrasound imaging is non-invasive, non-magnetizing and non-ionizing, offers high resolution, and is highly portable, thus offering significant clinical advantages.

Each color-Doppler sequence comprises a scalar field measured on a plane view over time. Nevertheless, all the ideas and algorithms presented in this work can be seamlessly ported to other flow imaging modalities, including multi-dimensional vector fields defined over volumetric regions of interest.

We defined *cohort-representative* LV flow fields by mapping the measured velocity data into a unified spatio-temporal mesh and averaging across patients from the same cohort. Previous echocardiographic and MRI studies have quantified LV flow metrics in healthy and diseased cohorts (Bermejo et al., 2014; Elbaz et al., 2014; Elbaz et al., 2017; Martinez-Legazpi et al., 2014). Most of these works report metrics averaged in space and time (e.g., flow KE averaged inside the LV and throughout the cardiac cycle) or choose specific regions and instants of time to pool data from different patients in the same cohort (e.g., residual volume at aortic valve opening) (Eriksson et al., 2013). Compared to these works, the spatio-temporal flow field alignment introduced in this study is novel and allows for visualizing, comparing and analyzing LV flow patterns with higher detail, both on an individual subject-by-subject basis and by groups. We ensured that the process of averaging across cohort patients did not significantly alter the resulting LV flow fields by comparing each *cohort-representative* flow field with the cohort’s medoid flow field, i.e., the flow from one individual in each cohort that is overall most similar to the flows from all members of the same cohort.

We applied POD to the *cohort-representative* LV flow fields and built ROMs for healthy subjects and patients with DCM and HCM. To enhance the interpretability of our results, we also performed VFM to the color-Doppler acquisitions and independently applied POD to the VFM data to build vectorial ROMs of LV flow. In both the color-Doppler and VFM data and in all three cohorts considered, the two highest-ranked modes accounted for >97% of the flow’s KE, capturing the filling/emptying jets (mode 1) and the diastolic vortex (mode 2). The eigenmode velocity vector maps exhibited appreciable differences among groups. Most notably, the DCM patients had shallower inflow/outflow jets that did not permeate the apical region but had large diastolic vortices that occupied the whole LV chamber. In contrast, the diastolic vortex was weak and confined at the LV base in HCM patients. While these results are broadly consistent with the existing literature, we note that LV flow ROMs provide a novel, data-driven approach to quantitatively synthesize each cohort’s representative flow features.

To test whether POD-based LV flow ROMs provide metrics that discriminate phenotypic differences, we used these ROMs to build three simple LV flow classifiers with different levels of supervision: supervised, weakly supervised, and quasi-unsupervised. The supervised classifier required training consisting of determining the *cohort-representative* LV flow fields and their POD-based ROMs. Then, each test patient was projected onto the three *cohort-representative* ROMs, the model residuals were calculated, and the differences between residuals were used to classify patients. The rationale was that, if a patient belongs to cohort *j*, the cohort-*j* ROM should perform better than the two alternative ROMs at reproducing the patient’s flow. This classifier performed favorably, with AUCs exceeding 0.75 and going up to 0.93. The weakly supervised classifier discriminated patients based on the ratio between the two highest-ranked patient-specific POD eigenvalues *V*2*J* = *B*_*2*_ / *B*_1_, i.e., the flow KE contained in the diastolic vortex normalized with the flow KE in the inflow/outflow jets, or in short, the vortex to jet ratio *(V2J)*. The only supervision in this second approach involves defining the *V2J* thresholds that separate the cohorts. The resulting AUCs, in the range 0.81 - 0.96 for PODmc, suggested that this weakly supervised classifier can outperform the supervised one for small and moderate training sample sizes. Semi-unsupervised classification leveraged classic clustering methods like mixed-Gaussian models or k-means clustering applied to the *B* _*α*_ (*α* = 1, …,*r*) multiplet obtained from patient-specific POD. It only involved specifying the number of ROM dimensions (*r*) and the number of clusters (three in our case, to account for the Control, DCM, and HCM cohorts). This approach achieved maximum accuracy for *r*≥4 and, similar to the other two methods, worked best in identifying DCM patients while it had most trouble distinguishing HCM patients from healthy controls. This differential performance could be caused by LV flow patterns sharing more features in the HCM and Control cohorts as compared to DCM, but we also note that our HCM cohort had the smallest sample size, whereas the DCM cohort had the largest.

Our analyses suggest that a one-dimensional LV flow ROM, i.e., a single numerical index per patient, contains sufficient information to discriminate between Control, DCM and HCM subjects, even after blinding the algorithm to LV chamber shape and size. This result is interesting because quantities like KE, dissipation or vorticity also provide a single numerical index when averaged over the whole LV chamber. Pathline analysis can be understood as a four-dimensional ROM since it describes LV flow using a four-component metric comprising direct flow, retained inflow, delayed ejection, and residual volume. Seen under this light, the discriminating power of POD is noteworthy. For reference, running LDA only on the global LV flow KE for our study cohort yields AUCs of 0.68, 0.69, and 0.52 for the binary classification problems DCM vs. HCM, DCM vs. Control, and HCM vs. Control, respectively. These accuracies are markedly inferior to the values of AUC = 0.96, 0.90, and 0.82 obtained when running LDA on *V*2*J*. Moreover, the metric *V*2*J* is no less interpretable than total KE. It is important to note that POD-based metrics were chosen data-drivenly to maximize discriminating accuracy, while they rely on the sparsity (rapid decay of mode energy with mode rank) and orthogonality (independence of eigenmodes) of POD for interpretability. On the other hand, classic hemodynamic metrics like KE are not data-driven and are chosen based on interpretability alone, as they can be related to physical principles.

Finally, it is worth underlining that we built ROMs and ran the classification pipelines to both “1D raw” color-Doppler maps the 2D vector map sequences obtained by VFM. Since VFM uses physical constraints to infer the cross-beam velocity component from the Doppler velocity, the color-Doppler and VFM datasets contained similar information. Therefore, there were little differences in classification performance from these two modalities. While visualizing the ROMs by VFM can make it easier to understand each cohort’s distinctive LV flow features, additional postprocessing of color-Doppler data is not necessary to obtain accurate classification based on LV flow, a desirable trait from the point of view of clinical translation.

### 4.1 LIMITATIONS AND FUTURE WORK

The dataset used in this study comprised 209 patient-specific, full-LV color-Doppler echocardiograms, each containing an average of 200x75 spatial pixels and 200 temporal frames. Given the moderate size of this database, hyperparameter tuning did not employ independent subgroups but was carried out as part of the training. To palliate the overfitting risk of this approach, we incorporated information penalties promoting low-order ROMs, performed 10-fold cross-validation, and adopted low-order discriminant functions for classification.

By design, we applied our methodology to three very distinct patient groups. Patients with HCM and DCM had fully manifested phenotypes and were easily distinguishable by standard clinical methods. While this selection provided a robust benchmark to derive LV flow metrics that can discriminate patients, the present paradigm could have broader applications and could be clinically useful in identifying more subtle phenotype variations. For example, characterization of flow patterns using this methodology could identify patients in the early stages of their disease before other structural changes become apparent. In addition, rigorous characterization of flow patterns using these techniques may serve as a useful biomarker to risk stratify patients within a disease group and serve as a useful surrogate endpoint for treatments.

We pre-processed each echocardiographic frame so that the time-varying endocardial border delineating the LV chamber and the color-Doppler map inside the chamber were mapped into a unit rectangle. The resulting flow maps were uninformative of ventricle size, endocardial morphology, or myocardial kinetics, all factors associated with cardiac health (Vigneault et al., 2019). The purpose of this reductionist approach was to test the value of LV flow patterns as a biomarkers of cardiac health without extrinsic biases. Certainly, the clinical deployment of ML tools based on LV flow patterns should be done in concert with existing methods for structural and kinetic analyses (Ferdian et al., 2020).

We used deliberately simple, well-established dimensionality reduction and classification methods. While the burgeoning literature in ML abounds with more sophisticate algorithms, our goal was to demonstrate that each subject’s LV flow signature is robust enough for uninvolved methods to quantify it and exploit it for patient phenotyping. This approach has the additional advantages of not requiring large training datasets and being computationally inexpensive. A thorough review of advanced ML models for LV flow pattern analysis would be beyond this manuscript’s scope but we can outline a few natural extensions of this study. Deep learning could be applied to LV flow fields for disease identification, similar to reported analyses of structural images (Tromp et al., 2022; Wehbe et al., 2023) . Contrastive POD (cPOD) could remove background LV flow features shared by healthy and diseased subjects, potentially improving cohort separability or identify subpopulations in DCM or HCM (Abid et al., 2018). Further work could also include exploiting multiple echocardiographic views, extending this methodology to other imaging modalities like, e.g., 4D flow MRI, and applying it to other cardiac chambers and disease conditions.

## 5. CONCLUSION

Reduced-order models of patient-specific echocardiographic measurements of left-ventricular flow provide an interpretable, low-dimensional representation of these flows that can be used to define novel metrics with clear clinical potential. The significance of these low-dimensional metrics is assessed by testing their ability to classify subjects with different cardiac pathologies. Their derivation can be tailored to the available training data and expert supervision workforce. This work demonstrates these ideas by building LV flow cohort-representative reduced-order models and classifiers for three patient groups: healthy individuals, patients with dilated cardiomyopathy, and patients with hypertrophic cardiomyopathy. The favorable classification accuracy, obtained with overtly uninvolved machine learning methods and using clinically accessible input data, suggests a novel strategy to realize the untapped potential of non-invasive cardiac flow imaging in the early detection and management of heart failure, as well as the development of personalized treatment strategies. Further research is needed to validate these findings and to investigate the clinical utility of flow-based biomarkers in conjunction with other metrics and in larger, more diverse patient populations.

## Data Availability

All data produced in the present study are available upon reasonable request to the authors

