## Supplemental Materials for "Deriving Explainable Metrics of Left Ventricular Flow by Reduced-Order Modeling and Classification"

María Guadalupe Borja, MEng^1^

Pablo Martinez-Legazpi, MEng, PhD^2^

Cathleen Nguyen, MEng^3^

Oscar Flores, AEng, PhD^14^

Andrew M. Kahn, MD^5^

Javier Bermejo, MD, PhD, FESC^6^

Juan C. del Álamo, AEng, PhD^1,3,7,8,*^

From the:

^1^Department of Mechanical and Aerospace Engineering, University of California San Diego, La Jolla, CA;

^2^Department of Mathematical Physics and Fluids, Facultad de Ciencias, Universidad Nacional de Educación a Distancia, UNED and CIBERCV, Madrid, Spain;

^3^Mechanical Engineering Department; University of Washington, Seattle, WA

^4^Department of Aerospace Engineering, Universidad Carlos III de Madrid, Leganés, Spain

^5^Division of Cardiovascular Medicine, University of California San Diego, La Jolla, CA

^6^Department of Cardiology, Hospital General Universitario Gregorio Marañón; Facultad de Medicina, Universidad Complutense de Madrid, Instituto de Investigación Sanitaria Gregorio Marañón and CIBERCV, Madrid, Spain;

^7^Center for Cardiovascular Biology; University of Washington, Seattle, WA.

^8^Division of Cardiology, University of Washington, Seattle, WA.

**SUPPLEMENTAL MATERIAL**

### SUPPLEMENTAL FIGURES

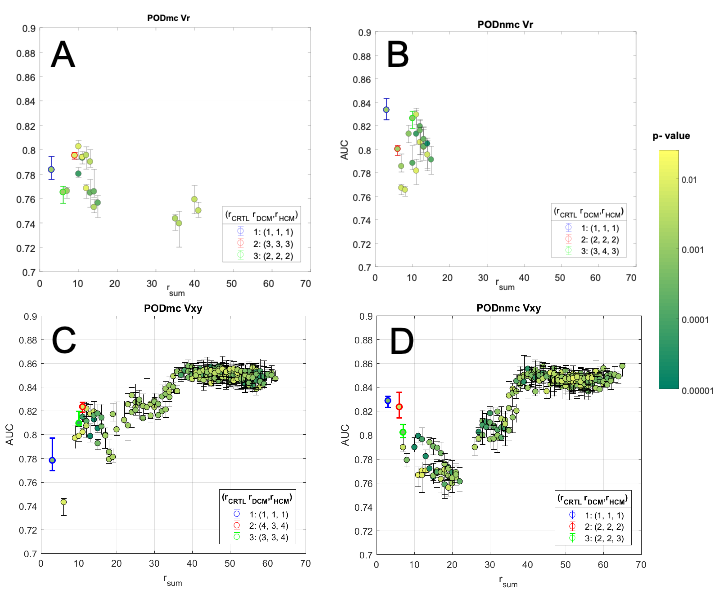

**SUPPLEMENTAL FIGURE 1**: Hyperparameter Grid Search. Classifier performance (AUC ROC, y-axis) vs. the sum of the number of modes in each cohort: r_sum_= r_ctr_+r_dcm_+r_hcm_ (x axis). Grid search was performed using as inputs both color-Doppler maps, *V_r_* (**Panels A&B**) and VFM maps obtained from color-Doppler, *Vxy* **(Panels C&D).** Results for classification based on mean centered POD **(Panels A&C)** and non-mean centered POD **(Panels B&D)** are presented.

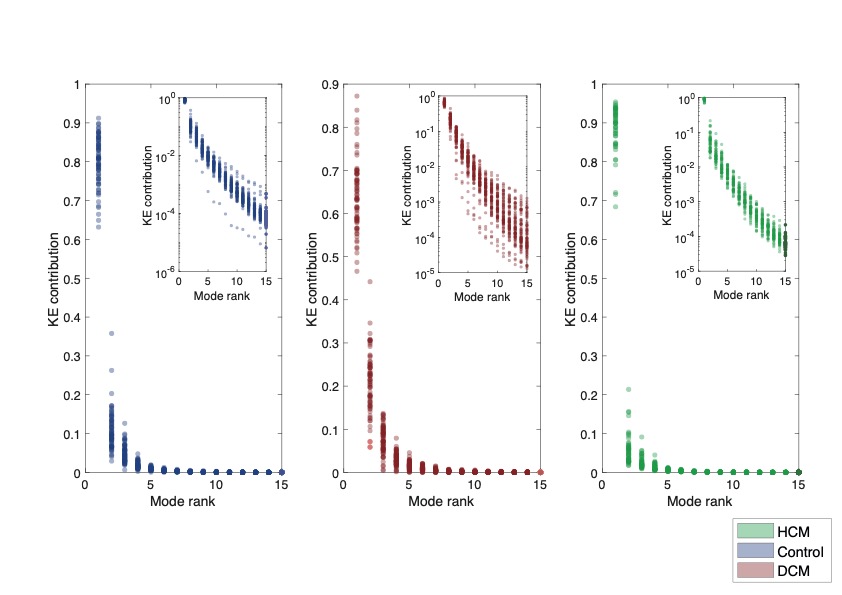

**SUPPLEMENTAL FIGURE 2**: KE content of POD eigenmodes for each cohort: Control subjects, (left) patients with dilated cardiomyopathy (center) and patients with hypertrophic cardiomyopathy (right). Inserts account for logarithmic plots.

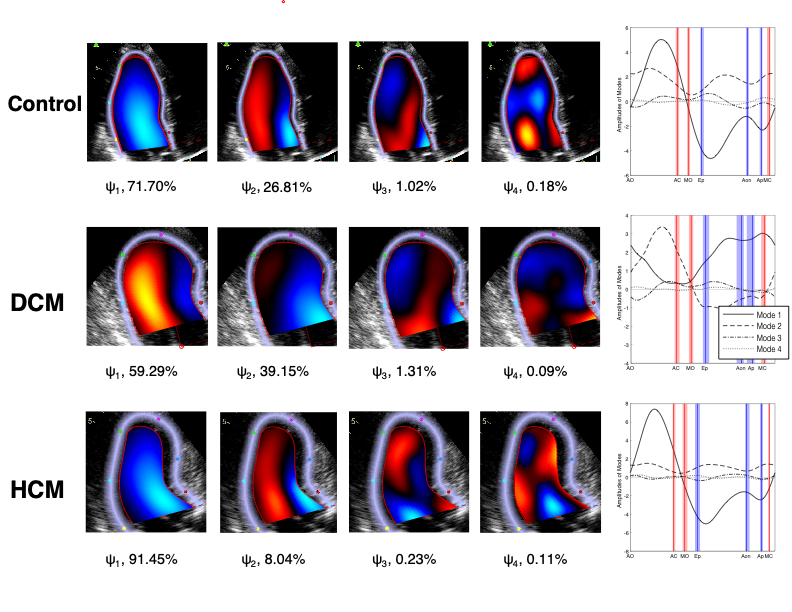

**SUPPLEMENTAL FIGURE 3**: Non-Mean centered POD modal decomposition of cohort-representative intraventricular, color-Doppler flow maps. Flow maps and POD were calculated on the rectangular unified spatio-temporal reference system, then remapped into a patient-specific anatomy from a randomly chosen patient within its cohort. The top, middle, and bottom rows indicate the Control, DCM and HCM cohorts, respectively. Columns 1^st^ to 4^th^ represent the four highest-ranked POD eigmode spatial maps ($\psi_{l}^{cohort}(x,y), l=1\cdots4$), whereas the 5^th^ column depicts these modes' time-dependent KEs (${b^{cohort}}_{l}(t)$, $, l=1\cdots4$). Red/blue lines represent each cohort's median event times: Aortic Valve Open (AVO), Aortic valve closure (AVC), E-wave onset (MVO), peak E-wave (Ep), A-wave onset (Aon) peak A-wave (Ap) and mitral valve closing (MVC). Shaded red and blue areas are the bootstrap median 95% confidence interval for each cohort.

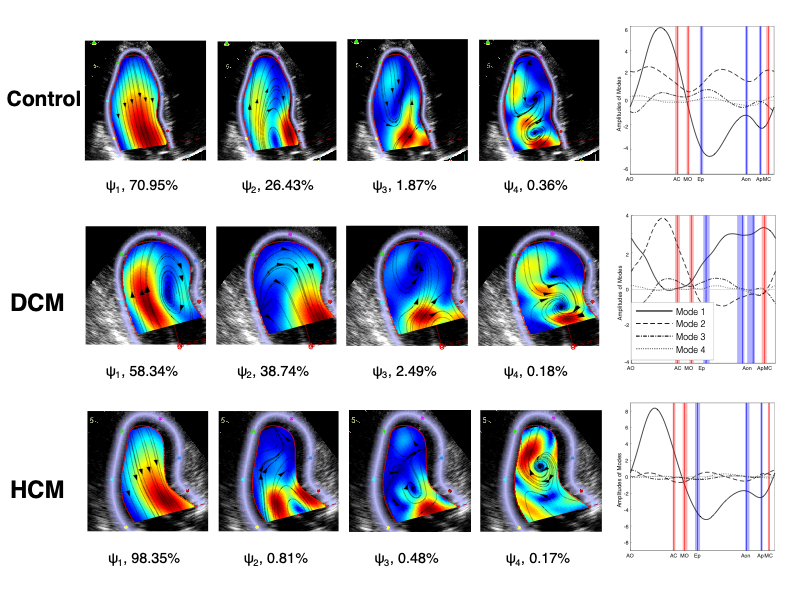

**SUPPLEMENTAL FIGURE 4**: Non-Mean centered POD modal decomposition of cohort-representative intraventricular velocity fields obtained by vector flow mapping (VFM). Each velocity component and its POD were calculated on the rectangular unified spatio-temporal reference system, then remapped into a patient-specific anatomy from a randomly chosen patient within its cohort. The top, middle, and bottom rows indicate the Control, DCM and HCM cohorts, respectively. Columns 1^st^ to 4^th^ represent the four highest-ranked POD eigmode spatial maps ($\psi_{l}^{cohort}(x,y), l=1\cdots4$), whereas the 5^th^ column depicts these modes' time-dependent KEs (${b^{cohort}}_{l}(t)$, $, l=1\cdots4$). Red/blue lines represent each cohort's median event times: Aortic Valve Open (AVO), Aortic valve closure (AVC), E-wave onset (MVO), peak E-wave (Ep), A-wave onset (Aon) peak A-wave (Ap) and mitral valve closing (MVC). Shaded red and blue areas are the bootstrap median 95% confidence interval for each cohort.

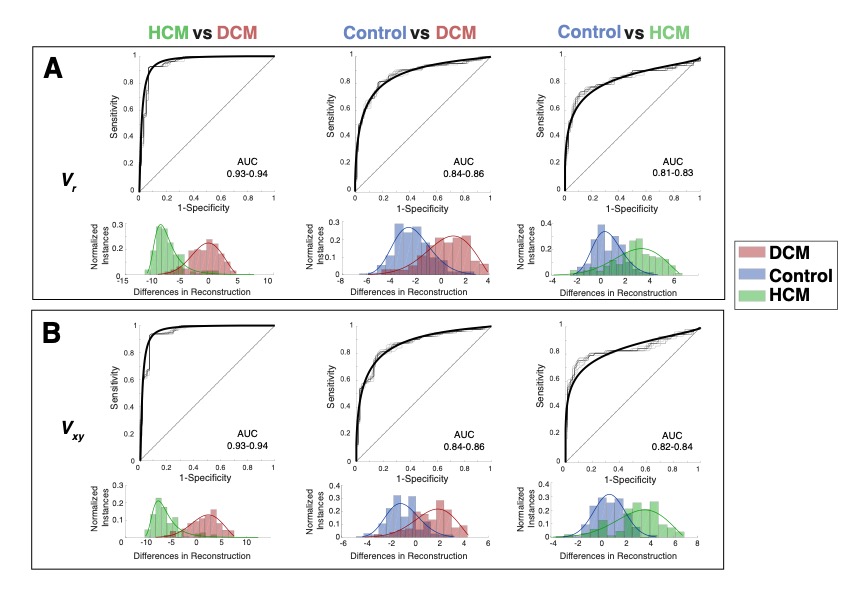

**SUPPLEMENTAL FIGURE 5**: Performance supervised classification based on the residuals of cohort-representative reduced-order models of LV using non-mean centered POD. **Panel A**: Model performance using color-Doppler maps as input data. **Panel B**: Model performance using VFM velocity maps as input data. From left to right, each column shows ROC curves (mean and repeated k-fold CV calculations) and residual histograms for the binary comparisons between HCM (green) & DCM (red), Control (blue) & DCM, and Control & DCM.

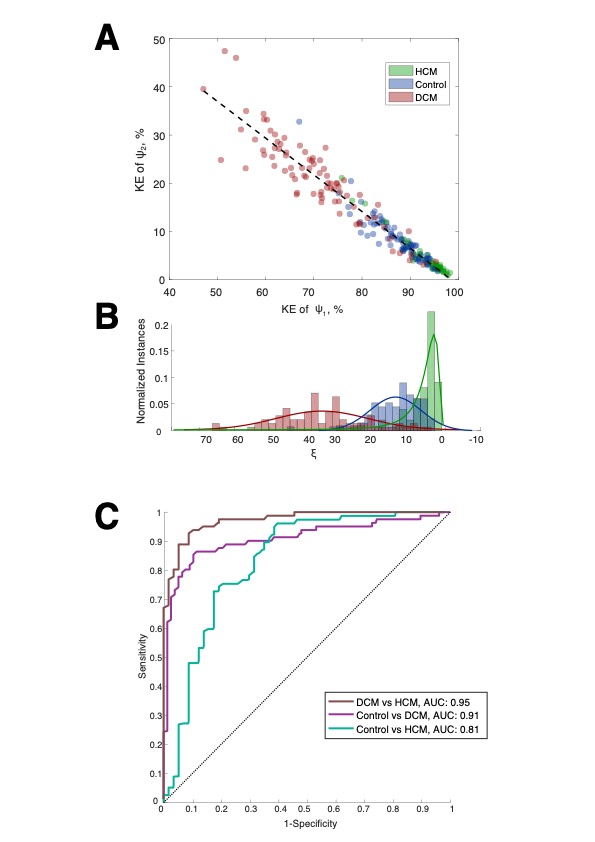

**SUPPLEMENTAL FIGURE 6**: Performance of weakly-supervised classifier based on patient-specific mean-centered POD using VFM velocity fields as input data. **Panel A**: Scatter plot of $B_{1j}$ and $B_{2j}$; each study subject is a data point colored according to their cohort. **Panel B**: Histograms of distance along the discriminant axis for each cohort. **Panel C:** Model performance. ROC curves are shown for the binary classifications: 1) Brown line: HCM (green) vs. DCM (red), 2) Purple line: Control (blue) vs. DCM and 3) Dark green: VOL vs. DMC.

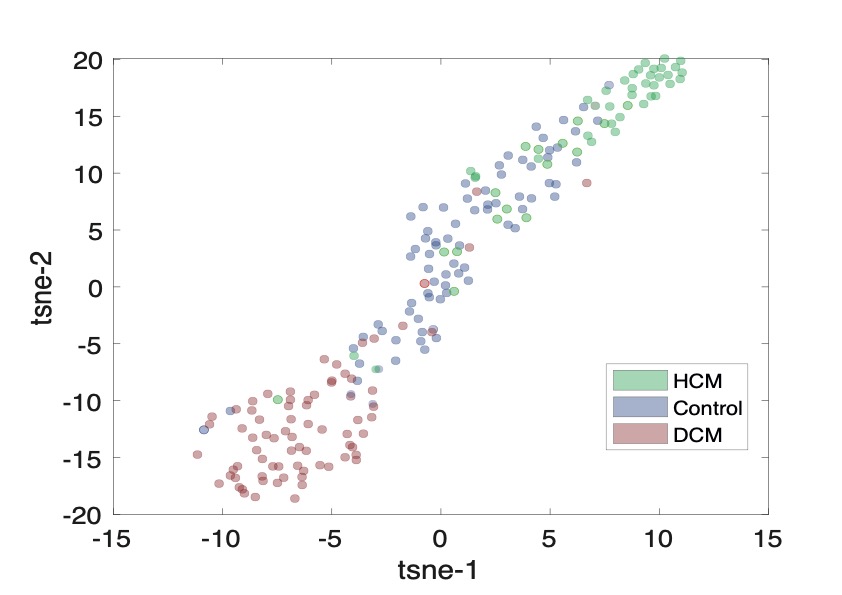
**SUPPLEMENTAL FIGURE 7**: Stochastic neighbor embedding (t-SNE) maps considering 15 dimensions, i.e., $B_{\alpha j} (\alpha=1,\cdots,r=15$), each study subject is a data point colored according to their cohort.

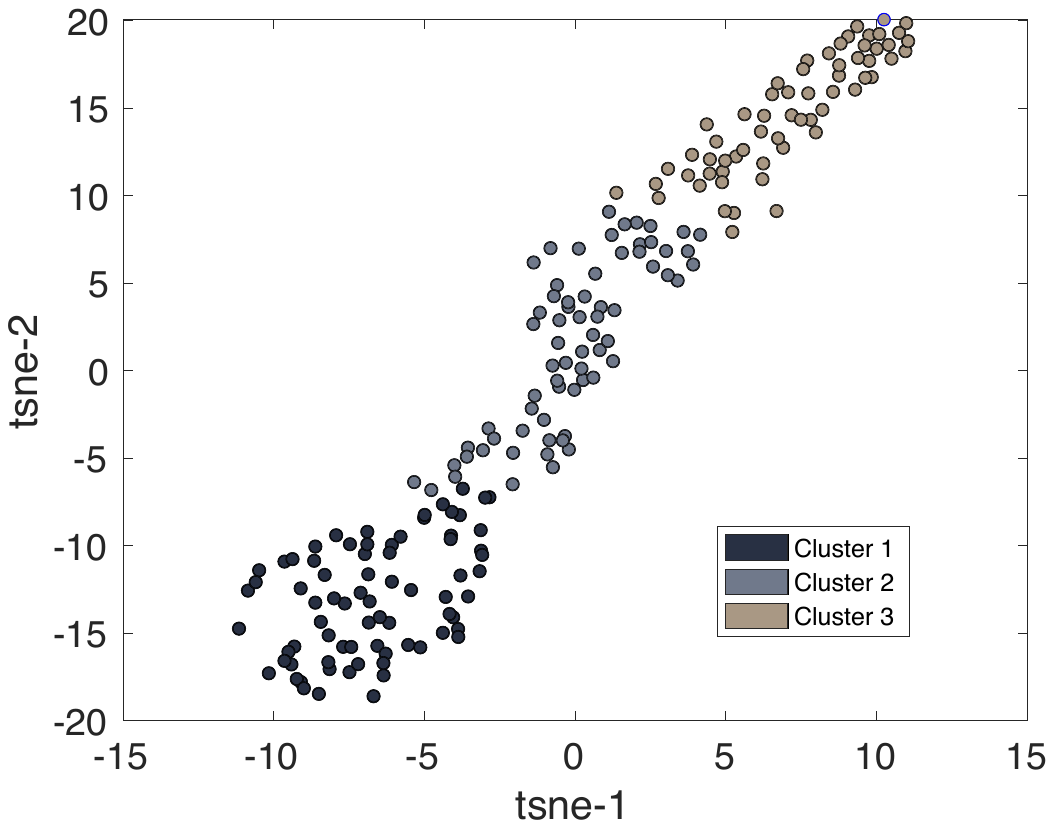

**SUPPLEMENTAL FIGURE 8**: Stochastic neighbor embedding (t-SNE) maps considering 15 dimensions, i.e., $B_{\alpha j} (\alpha=1,\cdots,r=15$), each study subject is a data point colored according to their cluster classification.
